# A mixed methods study of attitudes on location of gynaecological oncology outpatient care: a patient and healthcare professional questionnaire

**DOI:** 10.1101/2023.08.04.23293646

**Authors:** Rebecca Newhouse, Victoria Cullimore, Emily Hotton, Hilary Maxwell, Eleanor Jones, Jo Morrison

## Abstract

**Objective:** Gynaecological oncology place of care is often based on evolution of services, along historical professional boundaries, rather than user needs or preferences. We aimed to assess existing evidence, gather views of patients in the United Kingdom (UK) on their preferred place of outpatient care for gynaecological malignancies and evaluate alignment with preferences of healthcare professionals.

**Methods:** We performed a mixed methods study, including a scoping review, a patient survey, and a health care practitioner questionnaire. We collected quantitative and qualitative data, performing content analysis to determine current practice and impact on patients.

**Results:** We performed a mixed methods study, using a scoping review of the literature, patient survey and a healthcare practitioner (HCP) questionnaire, collecting quantitative and qualitative data. No studies were identified in our scoping review. We received responses from 159 patients and 54 gynaecological oncology HCPs. There was a strong preference for a dedicated gynaecological oncology setting (89% somewhat or very happy) (P<0.0001). 53% of patients were somewhat or very unhappy to have care co-located with general obstetrics and gynaecology services. Specifically, two key themes were identified through content analysis of qualitative data from patients: “environment and getting this right is vital”; and “our cancer should be the priority”. HCPs un-der-estimated the strong patient-preference to be seen in dedicated units. Of those who see patients within general obstetrics and gynaecology, only 50% said patients were seen at separate times/locations from obstetric patients.

**Conclusion:** This study demonstrates the significant impact of place of care on gynaecological oncology patients, which may be underestimated by HCPs.

**Key messages:** *What is already known on this topic:*

- Studies have shown that design of healthcare environment can significantly affect patient care, but have focussed on environmental factors, rather than co-location of services.

*What this study adds:*

- Gynaecological oncology patients indicated that co-location of clinics with general obstetrics and gynaecology was psychologically distressing or inappropriate, as they were at a different point in their life journey.
- Patients have a strong preference for their outpatient care to be provided in a dedicated gynaecologic oncology setting, away from women and children’s services.
- However, gynaecologic oncology services were frequently co-located with general obstetrics and gynaecology services, reflecting evolution of the subspeciality and service development, rather than patient need.

*How this study might affect research, practice or policy:*

- It is important to advocate for gynaecological oncology patients, to ensure that healthcare service infrastructure is designed around patient need, not historical professional boundaries.

## Introduction

Gynaecological oncology outpatient care encompasses diagnosis, treatment and follow-up appointments accessed by patients. In the United Kingdom, physical location of services varies depending on organisation of resources within individual institutions. As the subspeciality of gynaecological oncology developed from general obstetrics and gynaecology, care may be delivered by healthcare professionals (HCPs) with other responsibilities within the wider speciality [1]. Historical overlap of staff and resources with obstetrics, gynaecology and paediatrics means gynaecological oncology care in the UK is frequently delivered alongside other women and children’s services. This may be similar in many countries.

Obstetrics and gynaecology encompass a wide variety of different areas of medicine, making the specialty one of the most diverse areas of practice [2]. Consequently, gynaecological oncology care in the UK National Health Service (NHS) is seldom delivered in localities centred on those being investigated, treated or followed-up for cancer. Treatment for a gynaecological cancer can result in loss of fertility, and this sense of loss can be strong for people of all ages [3]. Additionally, nulliparity or infertility is associated with development of some gynaecological malignancies [4], which may further add to the emotional significance of where care is delivered, even when the individual is well outside of child-bearing age. Female reproductive organs form a broader feminine individual perception and changes caused by surgery can leave a lasting feeling of loss [5]. Not all patients may identify as women [6], which has implications for equity of access to care, especially if treated in a ‘women and children’s’ setting [7, 8].

Healthcare service design that is insensitive to patients’ needs communicates a negative message and makes people feel they are a low priority within the system [9]. The place of care provision is important for fostering a sense of support [9]. Reducing stress on patients associated with their environment is likely to increase the quality of a consultation; research has found that an elevated distress level is associated with an increased willingness to leave the decision control to the physician [10].

The experience a person has of their care has been outlined by the National Health Service (NHS) England as one of the three key principles in delivering cancer care [11]. There are a multitude of considerations in designing healthcare services and it is widely acknowledged that a patient-centred environment is desirable [12]. Compassionate care is central to good quality care, and clinical environment is one of the largest factors in delivering this aim [13].

To design environments to facilitate patient-centred care, we need to understand their experiences and views. We wished to understand the views of patients, both current and potential service users, in order to inform re-design of local services, and ascertain the gap between need and current provision.

## Methods

### Scoping literature review

We performed a scoping literature review to inform local service re-design using broad search terms on the themes of “gynaecology” “oncology or cancer” “place or site or environment” “treatment” “patient or service user or participant” in a PubMed search including studies up to April 2022 (see Supplementary 1).

### Study design and enrolment

A nested mixed methods approach was used to gather views of patients and HCPs. We used qualitative data collected in the patient survey to help further understand the quantitative data responses.

Women and people at risk of gynaecological cancers were invited to participate in a web-based survey by UK-based Gynaecological Oncology charities via email and social media (Twitter) (GO Girls, Peaches Womb Cancer Trust, Ovacome and Ovarian Action) with a total following of approximately 7000 people. All responses were voluntary and anonymous. No personally identifying data were collected. The survey was shared nationally, to broaden our understanding and avoid skewed responses in favour of existing local service models. HCPs registered with the British Gynaecological Cancer Society (BGCS) were invited to participate in a web-based survey distributed to members via email. The society includes approximately 500 trainees, consultants, and nurses working in medical, clinical, and surgical Gynaecological Oncology, both centres and units. A response rate of 10-20% was expected based on historic membership surveys.

### Data collection and analysis

Both quantitative and qualitative data were collected via QR code links to web-based surveys (MS Forms). HCPs were invited to complete the survey, if they were registered with the BGCS.

Quantitative data on responder characteristics and current service provision were collected. The patient survey included age, parity, and clinical information such as if they’d been investigated or treated for a gynaecological malignancy. A 5-point Likert scale was used to assess how they would feel being cared for in different clinical areas. HCPs were asked a selection of binary and multiple-choice questions. A 5-point Likert scale was used to collect participants views on the current location of care services. Quantitative data were analysed with Microsoft Excel [14] and Prism [15] using Chi Square analysis.

An open survey question was included to allow participants to expand on their experiences and gain deeper insights into their views. Content analysis was used for analysis, using the key steps that have been well described in the literature [16]. It enabled the identification of any commonalities and differences across participant responses. Qualitative data analysis was supported by NVivo 12 (QSR International, Melbourne, Australia).

### Ethics

The Somerset NHS Trust research ethics committee deemed ethics approval was not required, after completion of the Health Research Authority decision tool (http://www.hra-decisiontools.org.uk/research/), as this was a service improvement project to be used to inform re-design of local services.

## Results

### Scoping literature review

An initial scoping search of published literature retrieved only a very small number of potentially relevant papers in this area. Of the 4556 results from the PubMed search, none related specifically to our area of interest (Supplementary 2 and Supplementary 3). One study investigated patient views on change in service for a one-stop postmenopausal bleeding investigation clinic [17]. Two articles described the effects of place of care in teenage cancer treatment [9, 18].

### Patient attitudes

#### Quantitative data

A total of 159 patient responses were received, of which 140 had been investigated for gynaecological malignancies. The median age of the respondents was within the 55-64-year age group (Figure 1). Of those who answered, 97/158 (60.4%) had had children. The majority (157/159; 98.7%) identified as women, with one identifying as non-binary and one preferring not to say. Most (142/159; 89.3%) had been investigated for a potential gynaecological malignancy and 140/158 (88.6%) had been diagnosed with a gynaecological malignancy, reflecting targeting of respondents via gynaecological cancer charity communications and their social media channels. The median time taken to complete the survey was 2 min 47 sec (range of 27 sec to 36 min 14 sec).

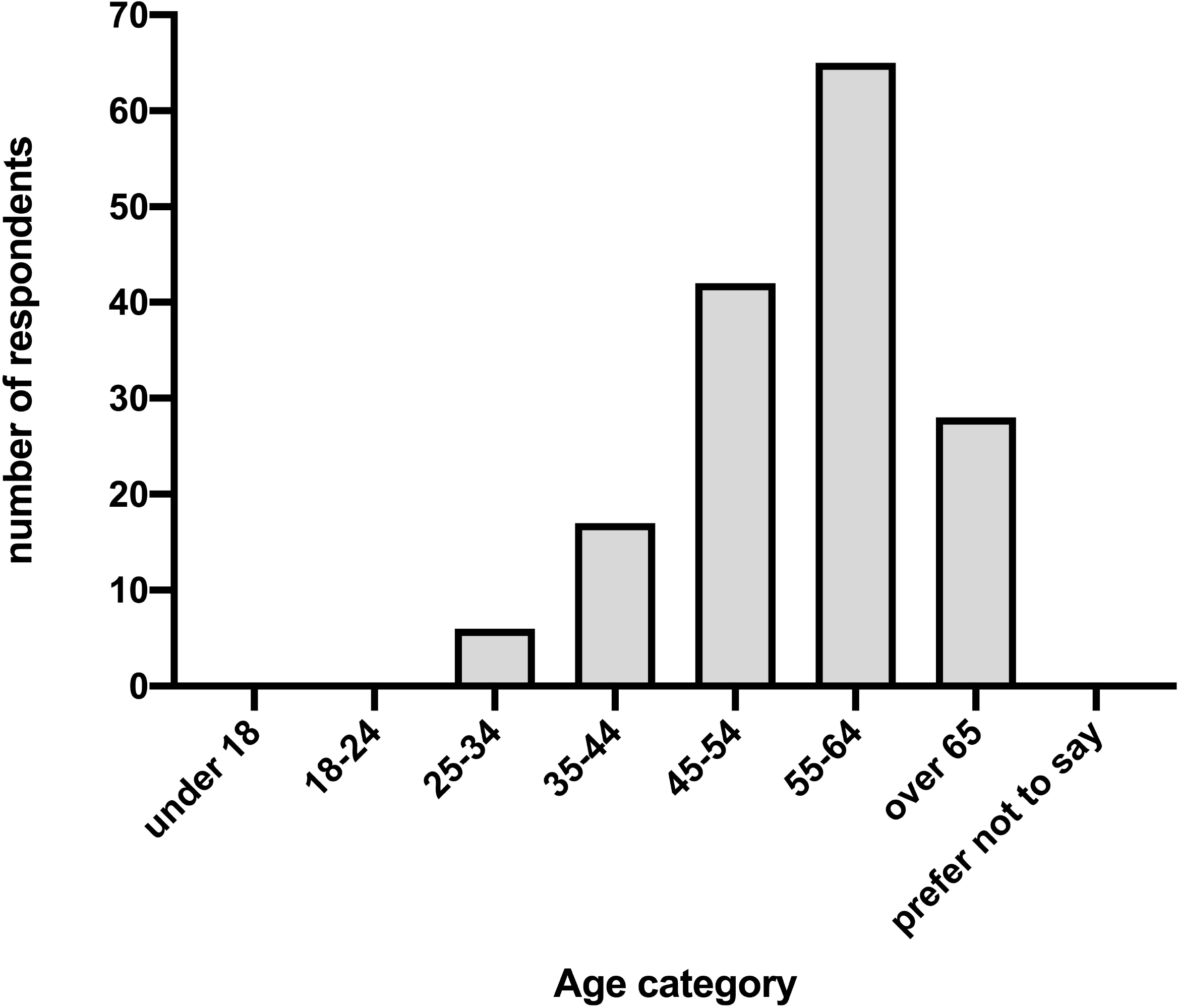

A gynaecological cancer/colposcopy unit was overwhelmingly the preferred place for care, with 89.4% being happy or very happy and only 4.9% somewhat or very unhappy with this option. The response to a mixed specialty cancer investigation unit was less positive, with 40.5% being somewhat or very unhappy and 33.6% neither happy nor unhappy. Over half of respondents (53.6%) were either unhappy or very unhappy to be seen within a general obstetrics and gynaecology department (Figure 2). There was significant difference between the options in terms of preference (P<0.0001).

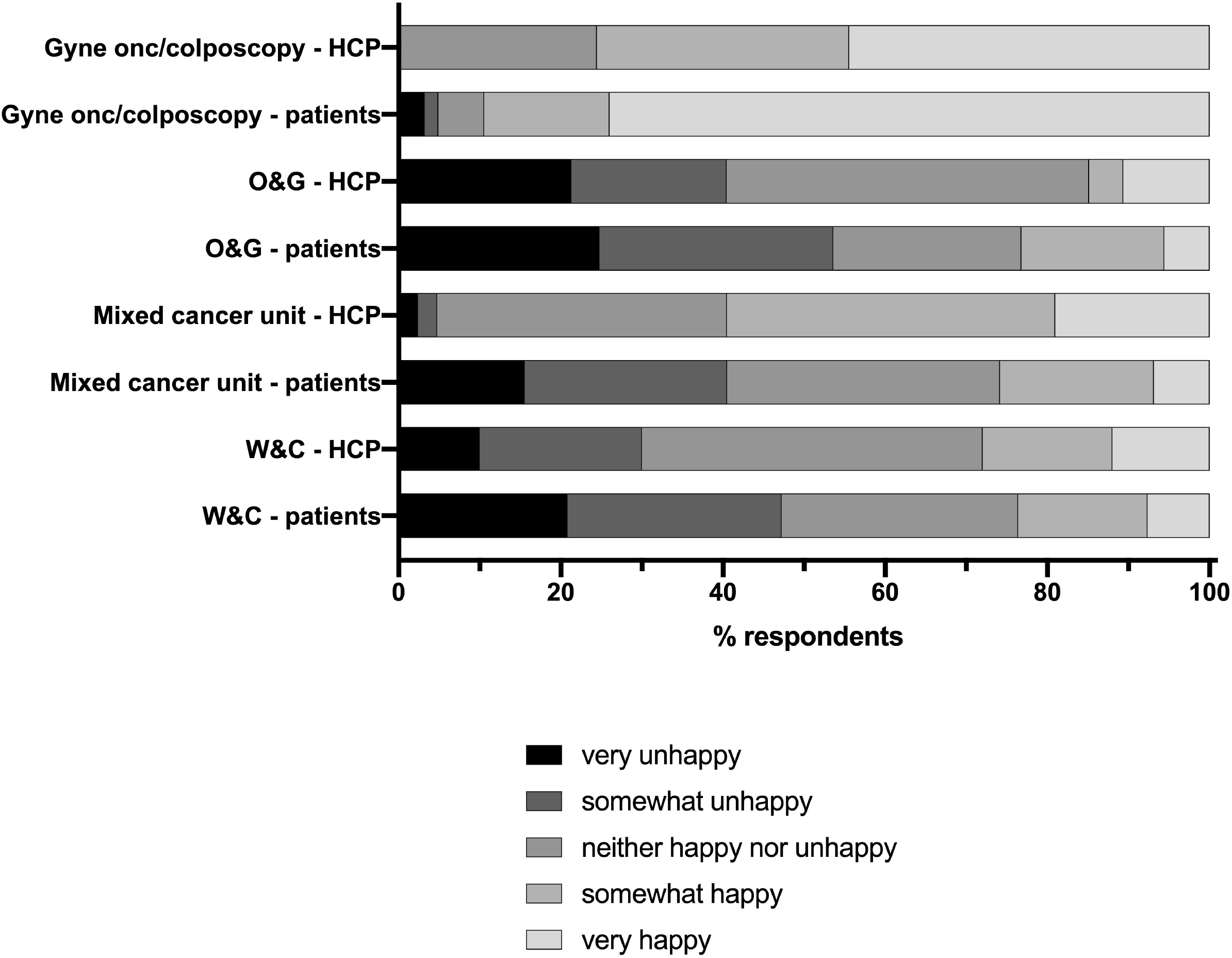

#### Qualitative data

One hundred and eight respondents provided free-text responses for analysis. Data are presented within the following themes: i) environment and getting this right is vital; and ii) our cancer should be the priority.

##### Theme one: Environment and getting this right is vital

Participants overwhelmingly considered the environment for their cancer care as a crucial consideration when determining their preference for location of care. The majority of respondents reflected on the insensitivity of having gynaecological cancer care in the same setting as maternity services:

> *P4: ‘The impact on fertility from cancer treatment, it’s emotive to be around pregnant patients or patients with young babies*.*’*

Many described their experiences of receiving care in a setting alongside maternity services as extremely negative, using descriptors such as: traumatic, insensitive, not appropriate, harrowing, thoughtless, stressful and upsetting. This was true for some women even if they had children of their own:

> *P33: ‘Walking through the maternity ward when you’re losing your last chance of being able to have a child is heart-breaking*.*’*
>
> There was an appreciation that attitudes of being seeing alongside maternity services may depend on individual circumstances of whether women have had children or not:
>
> *P21: ‘As a woman with children I would not mind where I where seen…Perhaps if I was younger and possibly finding out I would not be able to have children naturally being seen alongside maternity patients might be difficult*.*’*
>
> *P63: ‘It’s not fair - even as someone who never wanted children - to be lumped in with pregnant women*.*’*

Women also described an environment that was “happy” to be negatively triggering. This happiness was perceived to come from: seeing pregnant women, seeing children, seeing posters of babies and pregnant women or an awareness that other women in the waiting room were receiving positive pregnancy-related news:

> *P48: ‘If you’ve never had kids and/or want kids and then cancer is thrown in your path, the last thing you want to do is to be sat near happy pregnant women*.*’*
>
> *P75: ‘It was awful being in the same room as IVF patients, both of us upset as we watched pregnant women glow and being happy as we were receiving the worst news*.*’*

An additional perceived benefit to having care in a dedicated cancer setting was the notion of support: feeling supported by being surrounded by women who are going through the same experiences as each other, being in an environment that is felt to be safe, supportive and allowing women to feel at ease:

> *P10: ‘It is also sometimes comforting to know that others in the waiting room are there for broadly similar reasons*.*’*
>
> *P138: ‘I prefer being in clinic with people who understand what is happening’*

##### Theme two: Our cancer should be the priority

Table 1 demonstrates the views and attitudes of services users towards the gynaecological cancer diagnosis. There was a strong belief that care for gynaecological cancer should be delivered by specialist teams. Assumptions and beliefs included thoughts that clinicians: have greater interest, provide “better care” and more up-to-date. A small number of participants did reflect on personal positive experiences of mixed cancer centres:

**Table 1.**
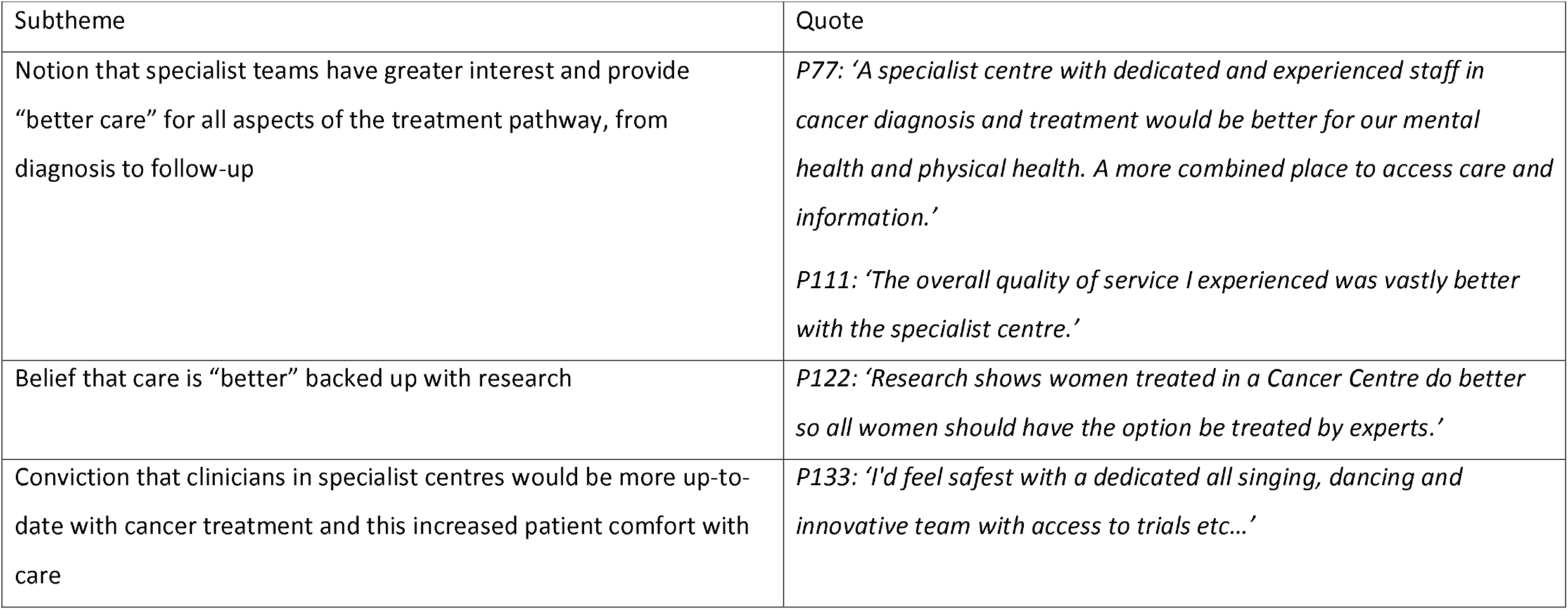
Theme two: “Our cancer should be the priority” - views and attitudes of patients towards the gynaecological cancer diagnosis.

> *P19: ‘I am being treated at a mixed speciality cancer centre and am extremely happy with my experience there*.*’*

### Healthcare professional attitudes

Fifty-four responses were received to the HCP survey from 41 separate institutions across the UK, representing 10% of the membership. The majority of respondents (81.5%) worked in gynaecological oncology; 13% in obstetrics and gynaecology. Just over half of respondents were consultants (53.7%) and 25.9% were specialist nurses. Two-thirds (66.8%) of respondents worked within cancer centres and 27.8% within cancer units; 3.7% stated “other”.

Investigation of patients referred with a suspected gynaecological malignancy is undertaken in a mix of settings: 35% are seen in a cancer investigations unit (31.4% gynaecological cancer/colposcopy, 3.7% mixed cancer investigation unit); 35.2% are seen in an obstetrics and gynaecology/women and children’s department. The remaining 29.6% selected “other” locations. Of those who saw patients within general obstetrics and gynaecology/women and children’s services, only 50% said patients were seen at separate times/locations from obstetric patients.

We compared the HCPs perceptions of patient preferences for each place of care and analysed with Chi Square analysis to see whether these differed from patient preferences for outpatient care location. HCPs were aware of the low level of acceptability to patients of being seen in a women and children’s unit (P = 0.224). However, HCPs over-estimated the acceptability of a mixed speciality cancer unit (P <0.0001) or a general obstetrics and gynaecology setting (P = 0.015), which were perceived by many patients as inappropriate places of care, and under-estimated the strong preference for a gynaecological cancer/colposcopy clinic (P = <0.0001) (Figure 3).

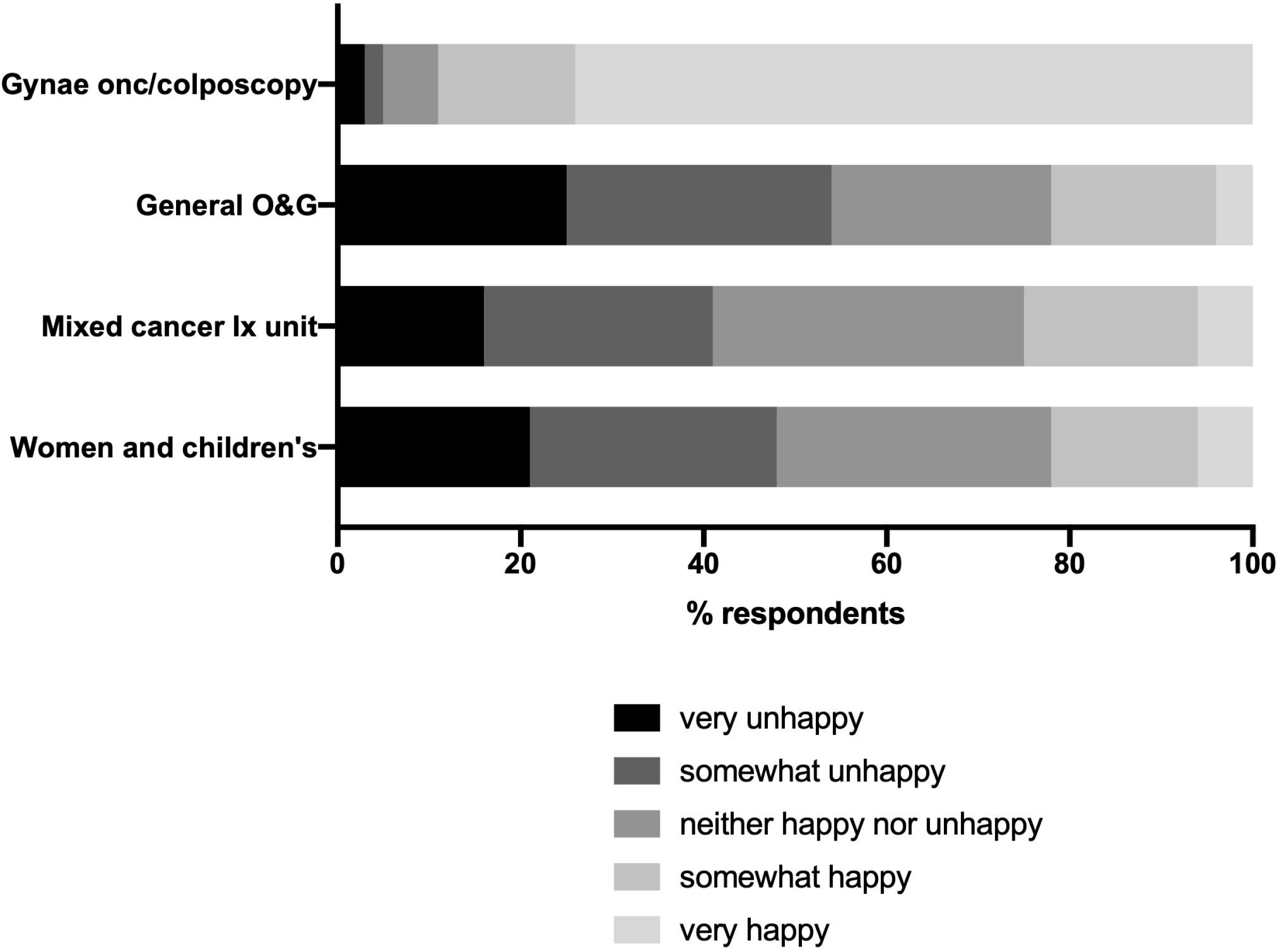

## Discussion

### Summary of Main Results

These findings highlight the significance of the care environment for those receiving investigations and treatment for gynaecological cancers. The responses from patients about their care experiences were emotive and highlighted potential ongoing harm to patients, if place of care is not considered.

Two themes were identified; the first “environment and getting this right is vital” and the second “our cancer should be the priority”. Respondents describe having their cancer care provided alongside maternity care as ‘traumatic’, ‘insensitive’, ‘not appropriate’, ‘harrowing’, ‘thoughtless’, ‘stressful’ and ‘upsetting’. These responses were not unique to those of childbearing age, as respondents were predominately no longer of childbearing age. Those that had no plans for future pregnancies were still affected by their shared environment. The emotional and psychological impact of gynaecological surgery is significant and long lasting [19], and can result in a feeling of having lost one’s femininity [20]. This ongoing impact highlights the importance of consideration towards place of care in the follow-up setting, as much as during diagnostics and treatment.

Some of the responses highlighted loss of fertility as a key concern with relation to those receiving care around them. Being in a ‘happy environment’ was also raised as being negatively triggering and laid bare the stark contrast between their own situation and that of pregnant women receiving positive pregnancy-related news around them. Posters of babies and pregnant women contributed to the ‘happy environment’ they were in, but felt estranged from.

A gynaecological cancer/colposcopy unit was found to be the preferred place for care in the quantitative data collected. This was reflected in the qualitative data where there was a notion of support when care was given in a dedicated cancer setting. Patients expressed feeling supported by being surrounded by people who are going through the same experiences as them, being in an environment that is felt to be safe and supportive and allowing people to feel at ease, reflected in the second theme “our cancer should be the priority” and supported by the quantitative data.

Only 18% of patients were happy or somewhat happy to receive care in a mixed speciality cancer investigations unit. Conversely, 45% of health care practitioners felt that the patients would be happy or somewhat happy in this mixed speciality cancer environment. This suggests that as clinicians we underestimate the importance to our patients of being in a specialist centre to their feeling of support.

### Results in the Context of Published Literature

These findings were reflected in the teenage cancer population, where Teenage Cancer Units are found to provide an appropriate environment where this specialist population feels comfortable and their needs are met. Being in a cancer unit dedicated to their care provided a supportive bond [9, 18]. It is interesting that, we consider the needs of teenagers with cancer, with associated research, but there is a dearth of information on this topic for gynaecological cancer care.

### Strengths and Weaknesses

It is important to consider the limitations of this study. There is a high risk of selection bias in the patient questionnaire. The invitation was shared by gynaecological oncology charities via social media (Twitter) and email newsletter. Therefore, participants may not be a representative sample of the intended population. Twitter users are more likely to be younger adults, urban dwellers and have a higher median income than the general population [21]. Additionally, those that chose to participate may have had particularly memorable experiences prompting them to respond. This could have resulted in greater extremes of views than those help by the target population. The professional views questionnaire was again open to selection bias. The majority of respondents (68%) work in cancer centres, broadly reflecting the membership of the BGCS. Responses are likely to have differed if a higher proportion of participants worked in cancer units, where care and facilities may be less specialist. It is therefore likely that nationally fewer patients are investigated for a potential gynaecological cancer or followed-up in a specialist setting.

### Implications for Practice and Future Research

These findings should shape future health care services, both in building design and how buildings are used. Although the opportunity to design a new hospital or department is rare, it is our responsibility to advocate for our patients in gynaecological oncology and ensure their experiences and preferences are heard. When services are being redesigned or relocated within an existing structure, these research findings should help us to consider if more compassionate care could be provided in an alternative location. Where care needs to be co-located with other services, attention should be paid to the environment and creating a sense of care around this group of patients. This could include dedicated entrances and waiting areas, considerate scheduling of clinics and timing of appointments, and mindfulness regarding the surrounding images and information.

## Data Availability

Data available on reasonable request from the authors.

## Acknowledgements

We thank:

Dr Andy Bryant for his help with statistical analysis.

BGCS council, especially Nick Wood and Deborah Lewis, for supporting the study and sharing the HCP questionnaire with the membership.

Anna Hudson (Ovacome), Georgina Tharp (Ovarian Action) and the teams at GO Girls and Peaches Womb Cancer Trust for distribution and promotion of the patient questionnaire.

Cate Newell and her team in the Somerset NHS Trust Library and Knowledge Service with their help and advice designing and running the literature search for the scoping review.

We are especially grateful to all those who shared their views and experience of accessing gynaecological oncology care and hope that this will help to design the services you deserve.

## Authorship

JM and RN developed the original idea for the study. RN and VC performed the scoping review. JM and RN designed the surveys with advice from HM, EJ, VC. HM, EJ, JM, RN, VC contributed to data collection. JM and RN performed quantitative data analysis; EH and RN performed qualitative analysis. RN, JM, VC, EJ, HM and EH contributed to writing and approved the final version of the paper.

## Data requests

Data available on reasonable request from the authors.

