## Supplementary 1 for "A mixed methods study of attitudes on location of gynaecological oncology outpatient care: a patient and healthcare professional questionnaire"

Search carried out in PubMed National Center for Biotechnical Information. Date of search 7/4/2022

1. Gynaecology/
2. (gynaecolog* or gynecolog*).tw.
3. or/1-2
4. Neoplasms/
5. (neoplasm or oncology or cancer*).tw.
6. or/4-5
7. Environment/
8. (plac* or site or environ*).tw.
9. or/7-8
10. Therapeutics/ or Therapy/
11. (therapeutics, or treatment* or therapy)
12. or/10-11
13. Patients/
14. (patient* or service* and user or participa*).tw.
15. or/13-14
16. 3 and 6 and 9 and 12 and 15

References from papers also searched.

gynaecologic OR gynecologic OR gynecologically OR gynecology [MeSH Terms] OR gynecology OR gynaecological OR gynecological

AND

neoplasms [MeSH Terms] OR neoplasms OR oncology OR oncology s OR cancer s OR cancerated OR canceration OR cancerization OR cancerized OR cancerous OR neoplasms [MeSH Terms] OR neoplasms OR cancer OR cancers

AND

place OR placed OR places OR placing OR placings OR site OR environ OR environment [MeSH Terms] OR environment OR environments OR environment s OR environs

AND

therapeutics [MeSH Terms] OR therapeutics OR treatments OR therapy [MeSH Subheading] OR therapy OR treatment OR treatment s

AND

patient s OR patients [MeSH Terms] OR patients OR patient OR patients s OR service OR service s OR serviced OR services OR services s OR servicing AND user OR participant OR participant s OR participants OR participate OR participated OR participates OR participating OR participation OR participations OR participative OR participator s OR participators
