## Supplementary figures and images for "A mixed methods study of attitudes on location of gynaecological oncology outpatient care: a patient and healthcare professional questionnaire"

### Supplementary 2

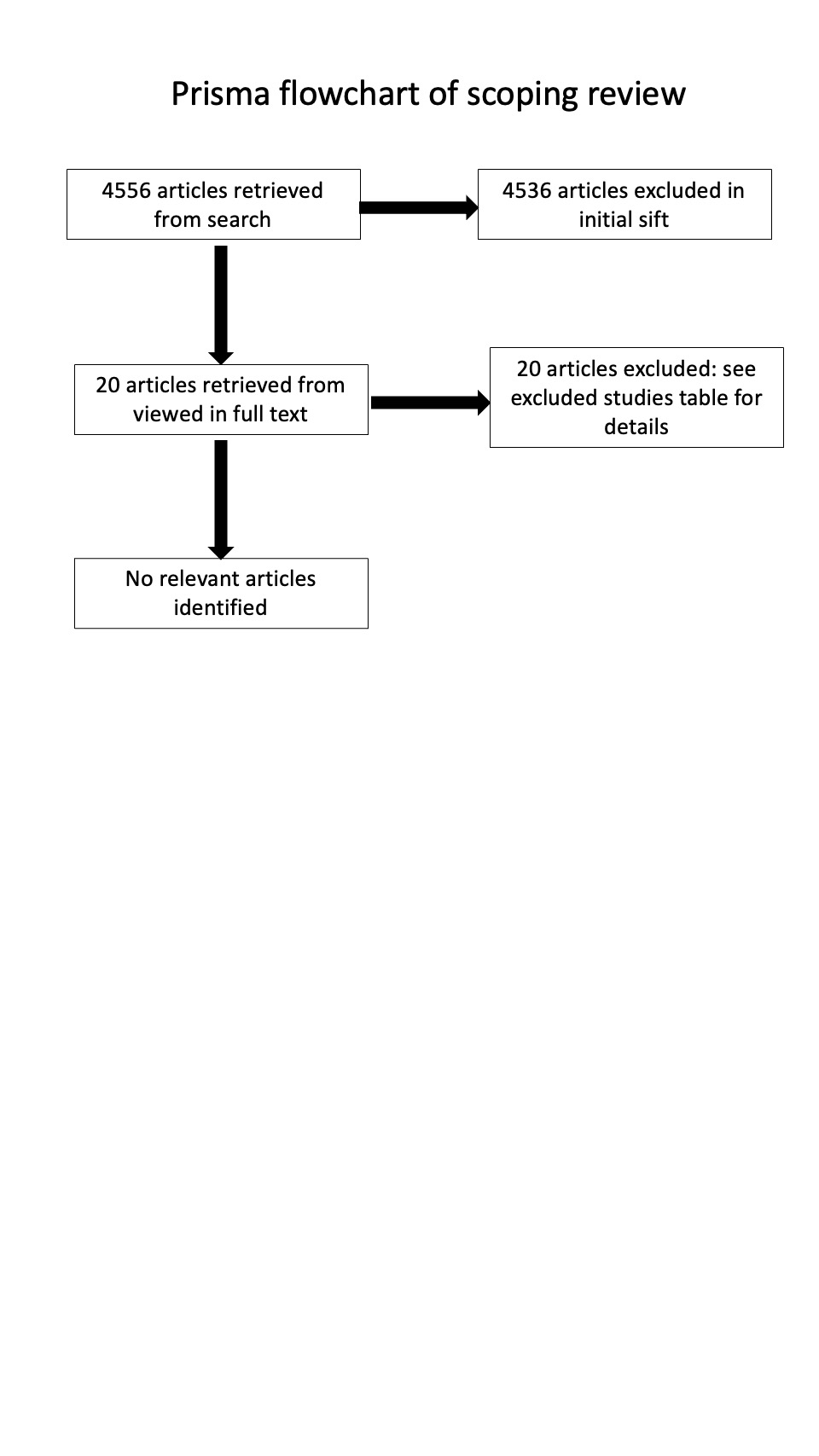
