## Supplementary 3 for "A mixed methods study of attitudes on location of gynaecological oncology outpatient care: a patient and healthcare professional questionnaire"

| Year published | Title | Authors | Purpose | Methods/data source | Results |
| --- | --- | --- | --- | --- | --- |
| Literature regarding the centralisation of gynae oncology care/structure of gynecologic oncology care | | | | | |
| 1999 | Improving outcomes in gynaecological cancer | Department of Health{Health, 1999 #30} | Guide commissioning, planning and development of gynaecological cancer services |  | Gynaecological cancer care should be centralised at a cancer centre, which is likely to improve survival and quality of life |
| 2012 | Centralisation of services for gynaecological cancer | Yin Ling Woo et al.{Woo, 2012 #14828} | Evaluate the centralisation of care for gynae oncology |  | Improved outcomes for women with gynaecological cancer in a cancer centre compared to non-specialist hospitals. Improved survival for ovarian cancer at centres. |
| 2011 | Improvements in survival of gynaecological cancer in the Anglia region of England: are these an effect of centralisation of care and use of multidisciplinary management? | Crawford and Greenberg{Crawford, 2012 #17619} | Evaluate the centralisation of care for gynae oncology in the Anglia cancer network |  | Significant stepwise improvement in survival of gynaecological cancers in 2000, coinciding with the major reorganisation of gynae-onc healthcare in the region as per the DOH report advised |
| 2015 | The optimal organisation of gynaecologic oncology services: a systematic review | Fung-Kee-Fung et al.{Fung-Kee-Fung, 2015 #17616} | Assess the relationship of the organisation of gynaecologic oncology services with patient survival and surgical outcomes. | Systematic review | Trend towards improved outcomes with centralisation of gynae-onc, particularly with advanced stage ovarian cancer. |
| 2009 | Variations in institutional infrastructure, physician specialization and experience, and outcome in ovarian cancer: a systematic review | du Bois et al.{du Bois, 2009 #17620} | Evaluate the impact of different physician and hospital characteristics on outcome in ovarian cancer patients | Systematic review | No impact of hospital type (teaching or non-teaching) on survival. Does not discuss individual hospital factors such as specific gynae onc units. |
| Literature regarding places of care/treatment | | | | | |
| 2007 | Change management in cancer care: a one-stop gynaecology clinic | Knight{Knight, 2007 #17607} | Description of how a DGH implemented a one-stop diagnostic clinic for suspected gynae cancer | Quality improvement | Discusses involving patients to plan services. Considering patient choice and convenience in services. |
| 1999 | Comparison of breast cancer patient satisfaction with follow-up in primary care versus specialist care: results from a randomised controlled trial | Grunfeld et al.{Grunfeld, 1999 #17622} | Assess patient satisfaction of transferring primary responsibility for follow-up of patients with breast cancer in remission from clinic to GP |  | Higher patient satisfaction with follow-up in general practice than hospital outpatients. When discussing follow-up with breast cancer patients, they should be provided with complete and accurate information about the goals, expectations, and limitations of the follow-up programme so that they can make an informed choice. |
| 2021 | Rapid Implementation of Telemedicine During the COVID‐19 Pandemic: Perspectives and Preferences of Patients with Cancer | Hasson et al.{Hasson, 2021 #17623} | Evaluate patients' perspectives and preferences regarding telemedicine and assess whether virtual communication affects the patient-physician relationship | Adult cancer patients who had had at least one successful telemedicine meeting were interviewed and completed patient satisfaction questionnaires | Patients did not feel that telemedicine appointments compromised medical care or the patient-physician relationship. |
| 2005 | A comparison of self-reported satisfaction between adolescents treated in a “teenage” unit with those treated in adult or paediatric units | Reynolds et al.{Reynolds, 2005 #17624} | Evaluate patient satisfaction for teenage cancer patients treated in two settings; first a split site unit (a paediatric ward and adult cancer centre in different locations), second a dedicated adolescent unit | Postal questionnaires of 65 adolescents who received cancer treatment between Sept 1997 and June 2000 | No significant difference in satisfaction of **overall care** between receiving treatment in a teenage cancer unit compared to an adult or paediatric unit. Significantly more satisfied with environmental aspects of care in teenage cancer unit. |
| 2003 | Where should teenagers with cancer be treated? | Whelan{Whelan, 2003 #17604} | Discusses place of care for treatment of teenage cancer patients |  | A teenage cancer unit provides an appropriate environment in which teenagers may feel comfortable and from which a multidisciplinary team can function. Some units cannot provide all aspects of care. |
| 2004 | A qualitative evaluation of an adolescent cancer unit | Mulhall, Kelly & Pearce{Mulhall, 2004 #17596} | An evaluation of the first specialist adolescent oncology unit in the UK | Semi-structured interviews of patients, parents and professionals | Adolescents with cancer's needs may be best met by specialist units. Being on a dedicated cancer unit compared to a general teenage unit provided a mutual sense of support and a place where cancer was not considered abnormal. |
| Literature regarding place of care in palliative care | | | | | |
| 2000 | Place of care in advanced cancer: a qualitative systematic literature review of patient preferences | Higginson, Sen-Gupta{Higginson, 2000 #17625} | Lit review of preferences for place of care and death among advanced cancer patients |  | Home is preferred location for most patients for place of death - mentions this as place of care in advanced cancer but more from a palliative perspective |
| 2008 | Preference for place of care and place of death in palliative care: are these different questions? | Agar, Currow et al.{Agar, 2008 #17626} | Research preferences of patients under palliative care for their place of care and place of death |  | "Place of care is not a euphemism for place of death" |
| 2018 | End of life care in Gynaecological Cancer | Kaushik{Kaushik, 2018 #17627} |  |  | Discusses variables related to place of death but nothing regarding place of care |
| Other relevant literature | | | | | |
| 2017 | Cancer patients’ control preferences in decision making and associations with patient-reported outcomes: a prospective study in an outpatient cancer center | Schuler et al.{Schuler, 2017 #17597} | To investigate possible determinants of the patient's decision control preferences with respect to patient characteristics and patient-reported outcomes | Self-administered electronic questionnaire | Higher age and higher levels of distress a/w "an increased willingness to leave the decision control to the physician". |
| 2021 | The hospital of the future: rethinking architectural design to enable new patient-centered treatment concepts | Amato et al.{Amato, 2022 #17599} | Proposing of a concept of "patient hub" in which care is "brought to the patient" | Simulation of workflows for patient hub and a traditional hospital | Multiple benefits in simulation of the patient hub (a completely patient-centred) model. |
| 2019 | Compassionate Care in Healthcare Systems: A Systematic Review | Tehranineshat et al. {Tehranineshat, 2019 #17598} | Determine the definition, fields, facilitating and inhibiting factors of compassionate care in healthcare systems and the interventions designed to promote it. | Systematic review | With regards to improving compassionate care, clinical environment and organisational values of the healthcare system are the largest facilitating and inhibiting factors |
| Literature regarding gynae patients experiences etc. | | | | | |
| 2016 | Woman experiencing gynecologic surgery: coping with the changes imposed by surgery | Silva and Vargens{Silva Cde, 2016 #17595} | Describe feelings and perceptions resulting from gynae surgery by women and analyse how they experience the changes caused by the surgery | Individual interviews | Two main themes: "perceiving a different body and feeling as a different person" and "building the meaning of mutilation." Different feelings of loss; actual loss e.g. oophorectomy (a physical loss) and then abstract re identity |
| 2008 | A qualitative study: beliefs and attitudes of women undergoing abdominal hysterectomy in Turkey | Reis et al.{Reis, 2008 #17601} | To reveal the beliefs and attitudes of women undergoing abdo hyst | Interviews | Five themes: (1) feminine identity, (2) husband/family relationships, (3) sexual life, (4) menopause, and (5) relatives’ opinions. Large long lasting impact of having an abdominal hysterectomy. |
| 2006 | The effect of hysterectomy on sexuality and psychological changes | Vomvolaki et al. {Vomvolaki, 2006 #17602} | Review of literature regarding the impact of hysterectomy | Literature review | Literature review on psychological effects of hysterectomy |

1. Department of Health. Improving Outcomes in Gynaecological Cancers. Guidance on Commissioning Cancer Services. Wetherby: Department of Health, 1999.

14. Kaushik S. End of Life Care in Gynaecological Cancer. Univeristy of Bristol, 2018.
