## Appendix 1 for "A mixed methods study of attitudes on location of gynaecological oncology outpatient care: a patient and healthcare professional questionnaire"

### Service user questionnaire

We are hoping to get your views about where you would prefer to be investigated for a suspected gynae cancer, so we can help design systems that are better for service users/patients. Currently services may be located within women and children's services or more general cancer services, and we understand that this may be challenging for some people. Thank you for your time and thoughts.

1. How old are you?

- ◯ Under 18
- ◯ 18-24
- ◯ 25-34
- ◯ 35-44
- ◯ 45-54
- ◯ 55-64
- ◯ over 65
- ◯ prefer not to say

2. Have you had any children?

- ◯ Yes
- ◯ No
- ◯ Prefer not to say

To attract more attention and response, we suggest adding an immersive theme style.

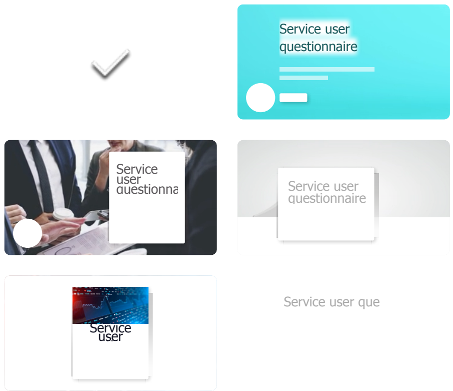


View all

Background music

3. Have you ever been investigated for a possible gynae cancer?

- ◯ Yes
- ◯ No
- ◯ prefer not to say

4. Have you ever been diagnosed with a gynae cancer or pre-cancer?

- ◯ Yes
- ◯ No
- ◯ prefer not to say

5. If you were investigated for a possible gynae cancer, how would you feel about being seen in the following departments?

| | very unhappy | Somewhat unhappy | Neither happy nor unhappy | Somewhat happy | Very happy |
|---|---|---|---|---|---|
| Women and children's department | ◯ | ◯ | ◯ | ◯ | ◯ |
| mixed specialty cancer investigation unit (mixed sex - combined with breast and bladder cancer patients) | ◯ | ◯ | ◯ | ◯ | ◯ |
| general obstetrics and gynae unit (combined with maternity services) | ◯ | ◯ | ◯ | ◯ | ◯ |
| dedicated gynae cancer/colpos copy unit | ◯ | ◯ | ◯ | ◯ | ◯ |

6. Where would your preferred place to be investigated for a possible gynae cancer out of the following?

- ◯ Women and children's department
- ◯ mixed specialty suspected cancer investigation unit
- ◯ general obstetrics and gynae unit (combined with maternity)
- ◯ dedicated gynae cancer/colposcopy unit
- ◯ no preference
- ◯ rather not say

7. If you have had treatment for a gynae cancer, where would you prefer to have your follow up appointments?

- ◯ Mixed specialty cancer centre
- ◯ Women and children's unit
- ◯ general obstetrics and gynae unit (combined with maternity)
- ◯ dedicated gynae cancer centre
- ◯ no preference
- ◯ prefer not to say

8. Please let us know the reasoning behind your preferences and any further comments

9. What gender do you identify as?

- ◯ Woman
- ◯ Man
- ◯ Non-binary
- ◯ Prefer not to say

This content is neither created nor endorsed by Microsoft. The data you submit will be sent to the form owner.

Microsoft Forms
