## Appendix 2 for "A mixed methods study of attitudes on location of gynaecological oncology outpatient care: a patient and healthcare professional questionnaire"

### Location of gynaecological oncology services

Having asked patients their views, we are conducting a national survey to find out where patients with suspected and proven gynaecological cancer are seen within our services.

We would be very grateful for your time in completing this short survey to inform us of the situation nationally. We aim to present the data, and will do so anonymously, but would be grateful if you could include your hospital site [illegible]

1. What is your job role?

- ◯ consultant
- ◯ clinical fellow
- ◯ specialist trainee
- ◯ CNS
- ◯ Subspecialist trainee
- ◯

2. What is your specialty?

- ◯ General obstetrics and gynaecology
- ◯ Gynaecology
- ◯ medical oncology
- ◯ Gynaecological Onoclogy
- ◯ clinical oncology
- ◯

To attract more attention and response, we suggest adding an immersive theme style.

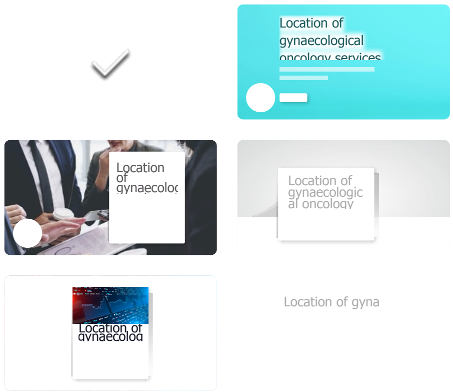


View all

Background music

3. What is the name of the site (Hospital and Trust, if different)

4. Is this site a Cancer centre or unit?

- ○ Centre
- ○ Unit
- ○ Other

5. Where do you see women being investigated for a suspected gynae cancer?

- ○ General obstetrics and gynaecology unit (combined with maternity services)
- ○ Dedicated gynae cancer/colposcopy unit
- ○ Mixed specialty cancer investigations unit
- ○ Women and children's department
- ○ Other

6. If you share space with O&G patients for diagnostic clinics, are the gynae onc clinics held at a time when obstetric patients/early pregnancy patients are using the same waiting area at the same time?

- ○ Yes
- ○ No
- ○ Maybe
- ○ Other

7. Where do you see women being followed up for a suspected gynae cancer?

- ◯ Dedicated gynae cancer/colposcopy unit
- ◯ General obstetrics and gynaecology unit (combined with maternity services)
- ◯ Mixed specialty cancer investigations unit
- ◯ Women and children's department
- ◯ Other

8. If you share space with O&G patients for follow up clinics, are the gynae onc clinics held at a time when no obstetric patients/early pregnancy patients are using the same waiting area at the same time?

- ◯ Yes
- ◯ No
- ◯ Maybe
- ◯ Other

9. Where are your gynae oncology surgical patients cared for post op?

- ◯ general surgical ward
- ◯ gynaecology ward separate to other O&G services
- ◯ gynaecology ward within general O&G services building
- ◯ Other

10. Where are your gynae onc theatres located

- ◯ within gynaecology theatres (no obstetrics)
- ◯ within general surgical theatres
- ◯ within general O&G department
- ◯ Other

11. How happy are you with the following settings to see patients in with suspected or confirmed gynaecological cancer?

| | Very unhappy | Somewhat unhappy | Neither happy nor unhappy | Somewhat happy | Very happy |
|---|---|---|---|---|---|
| Women and children's department | ◯ | ◯ | ◯ | ◯ | ◯ |
| Mixed specialty cancer investigations unit | ◯ | ◯ | ◯ | ◯ | ◯ |
| General obstetrics and gynaecology unit (combined with maternity services) | ◯ | ◯ | ◯ | ◯ | ◯ |
| Dedicated gynae cancer/colpos copy unit | ◯ | ◯ | ◯ | ◯ | ◯ |

12. How happy **DO YOU THINK YOUR PATIENTS ARE** with the following settings to see those with suspected or confirmed gynaecological cancer?

| | Very unhappy | Somewhat unhappy | Neither happy nor unhappy | Somewhat happy | Very happy |
|---|---|---|---|---|---|
| Women and children's department | ○ | ○ | ○ | ○ | ○ |
| Mixed specialty cancer investigations unit | ○ | ○ | ○ | ○ | ○ |
| General obstetrics and gynaecology unit (combined with maternity services) | ○ | ○ | ○ | ○ | ○ |
| Dedicated gynae cancer/colpos copy unit | ○ | ○ | ○ | ○ | ○ |

13. Please tell us more about your answer or any further comments

This content is neither created nor endorsed by Microsoft. The data you submit will be sent to the form owner.

Microsoft Forms
